# Characterizing Sleep Disorders in an Autism-Specific Collection of Electronic Health Records

**DOI:** 10.1101/2021.10.29.21265659

**Authors:** EV Singer, M Niarchou, A Maxwell-Horn, D Hucks, R Johnston, J Sutcliffe, LK Davis, BA Malow

## Abstract

**Background:** Sleep problems are common in individuals with autism spectrum disorder (ASD). This study reviews one detailed approach to querying the electronic health record (EHR).

**Methods:** We developed methods for identifying individuals with ASD and defined their sleep problems using International Classification of Diseases (ICD) codes or key words. We examined treatment responses to melatonin.

**Results:** Sleep problems were documented in 86% of patients and using specific keywords yielded more sleep diagnoses than ICD codes alone. About two-thirds of patients benefitted from melatonin.

**Conclusions:** Our study provides a framework for using deidentified medical records to characterize sleep, a common co-occurring condition, in ASD. Using specific keywords could be helpful in future work that queries the EHR.

## Background

ASD is a heterogenous disorder with core features of impairments in social communication and restricted behaviors (Kim et al., 2014). Sleep problems are highly prevalent in autism spectrum disorder (ASD) and contribute to challenging daytime behavior and family stress (Herrmann, 2016). Children and young adults with ASD have an increased frequency of sleep problems than their typically developing peers (Liu et al., 2006; Reynolds et al., 2019). Carmassi et. al. (2019) reviewed 74 studies reporting prevalence of sleep disturbance in ASD between 64% and 93%. There are a wide variety of sleep problems in ASD, with multifactorial causes. Insomnia is one of the most common sleep issues (Veatch et al., 2015). Insomnia is defined as difficulty initiating or maintaining sleep, with adequate opportunity to sleep, accompanied by daytime consequences (Sateia, 2014). Given their neurodevelopmental disability, the consequences of poor sleep, such as insomnia, may be more profound in individuals with ASD.

Prior studies of sleep problems in ASD have evaluated sleep objectively with polysomnography (PSG) or actigraphy, subjectively with validated questionnaires and sleep diaries completed by parents, or a combination of both (Carmassi et al., 2019). These forms of sleep evaluation have limitations with small sample sizes and are often cross-sectional rather than longitudinal. Also, they may only reflect the parental experience, which can be skewed and represent parents’ own perceptions of their child’s sleep disturbances. Furthermore, even objective data such as PSG have limitations. Some individuals with ASD cannot tolerate PSG, and PSG is subject to a first-night effect of decreased total sleep time and lower sleep efficiency (Malow et al., 2006). While actigraphy provides reasonable estimates of sleep parameters compared with PSG, it does not capture important aspects of sleep, such as bedtime resistance (Malow et al., 2016; Walia & Mehra, 2019). Collection of PSG or actigraphy data is costly, burdensome, and limits sample size. One way to characterize sleep problems in large datasets is to take advantage of electronic health records (EHR).

The EHR is a rich resource for characterizing medical conditions, including course over time and response to treatment. At our institution, we have access to a database of de-identified patient information that creates a mirror-image of the EHR. The EHR offers a more complete assessment of a patient’s health history with different clinical notes, communications with parents and medication records listed. This is particularly useful when characterizing complex conditions, such as ASD. The EHR’s advantages include a longitudinal database and a variety of information including past medical history, medication lists, laboratory results, clinic, and hospital documentation.

Prior sleep research in obstructive sleep apnea has utilized EHR by querying International Classification of Diseases (ICD) diagnostic codes (Hinkle & Kaelber, 2021; Keenan et al., 2020). However, identifying sleep problems within the EHR has not been well studied. Using this de-identified health record data, we aimed to assess sleep problems in patients with ASD. The purpose of the study was to determine the feasibility of a detailed chart review process within an EHR to identify patients with ASD and subsequently characterize their sleep problems. We first validated an algorithm for detecting ASD cases, using ICD diagnostic coding and chart review to classify patients, as in previous studies (Brooks et al., 2021; Bush et al., 2017). We then assessed sleep problems within the validated sample of individuals with ASD using chart review. We utilized a REDCap data form to build our database and generate reports. Here we describe our novel approach and discuss its benefits and challenges.

## Methods

To characterize sleep problems in ASD in our EHR, we first developed a standardized set of patients with ASD. General information on our de-identified EHR and how individuals with ASD were identified is presented below and followed by our process for characterizing sleep problems.

### Institutional Electronic Health Records (EHR) database

Approval was obtained through the Institutional Review Board to review the Synthetic Derivative (SD). The SD is a de-identified database of Vanderbilt University Medical Center’s EHR system that contains over 3 million unique individual records (Roden et al., 2008). The SD contains diagnostic and procedure codes, demographics, problem lists, medications, clinic and hospital notes and clinical communications. All charts in the SD had names redacted and dates shifted by up to 1 calendar year consistently within each record but differing across all records to provide anonymity. Dates are altered throughout the SD system as an additional layer of privacy protection. Individuals age at time of record review is available. However, patient ages in clinical notes are redacted and replaced with an age range, for example “birth-12 year old”.

### Identification of individuals with autism

In the SD, we generated a set of putative ASD cases for manual chart review by deploying an algorithm that required the presence of at least one occurrence of any of the following thirteen ICD codes (ICD 9: 299.0, 299.00, 299.01, 299, 299.8, 299.80, 299.81, 299.90, 299.91; ICD 10: F84.0, F84.8, F84.5, F84.9) AND the presence of at least one of the following keywords; “asperger”, “autism”, “autistic”, “pervasive developmental disorder”(See Table 1). With guidance from experts in the clinical presentation of ASD, we developed a chart review rubric to facilitate ascertainment of true cases within the SD set. The rubric allowed for the reviewers to stratify cases into low-evidence, medium-evidence, and high-evidence subsets. Cases were defined as follows: Low-evidence cases were required to have at least one affirmative mention of ASD in a chart of any type. Medium-evidence cases were distinguished by the presence of either a psychological evaluation form with an explicit ASD diagnosis OR at least two affirmative mentions of ASD by ASD-specific provider(s) (neurology, psychiatry, developmental pediatric, behavior therapy, occupational therapy, speech language pathology). High-evidence cases were distinguished by the presence of any of the following: a psychology evaluation with confirmatory Autism Diagnostic Observation Schedule, a clinic visit by an autism specialist, a clinic visit for medication management for ASD or two mentions of the patient being treated in Treatment and Research Institute for Autism Spectrum Disorder (TRIAD) clinic, even without the actual clinic note.

**Table 1:** Search criteria for defining ASD in the EHR

| Filter Documents by Keyword | Search ICD Codes |  | ASD Forms data |
| --- | --- | --- | --- |
| • Autism<br>• Autistic<br>• Asperger<br>• PDD | **<u>ICD-9 CODES</u>** | **<u>ICD-10 CODES</u>** | • TRIAD Autism Specialist<br>• Developmental Medicine and Cognition<br>• Developmental Pediatrics Evaluation<br>• Psychology Evaluation |
|  | 299.0 | F84.0 |  |
|  | 299.00 | F84.8 |  |
|  | 299.01 | F84.5 |  |
|  | 299 | F84.9 |  |
|  | 299.8 |  |  |
|  | 299.80 |  |  |
|  | 299.81 |  |  |
|  | 299.90 |  |  |
|  | 299.91 |  |  |

Chart reviewers were instructed to note the presence of the above criteria and to carefully evaluate the available information, making note of conflicting evidence if present, in reaching their determinations. Subjects were excluded if their records did not satisfy the minimum criteria for low-evidence cases, if their records included a psychology evaluation that did not support an ASD diagnosis, or if conflicting evidence placed a diagnosis in doubt.

An initial un-blinded training sample using 65 charts was performed to ensure the reviewers were correctly implementing the chart review rubric. After initial training, two chart reviewers performed a blinded review of a single set of 25 charts, and inter-rater reliability was estimated at 0.78 (with agreement on 24/25 charts) with regard to inclusion/exclusion. Next, the chart reviewers performed a blinded review of an additional 50 charts with perfect agreement regarding inclusion/exclusion. The remaining charts were divided between the reviewers and were reviewed independently. Each chart reviewer recorded his or her determinations in an Excel spreadsheet and included evidence copied from the record to justify his or her determinations. Upon conclusion of the review, an expert in ASD clinical presentation reviewed the evidence presented by the chart reviewers and confirmed that the evidence presented supported the determinations.

### Chart Selection

This review process generated over 800 patient charts with either low, medium, or high evidence of ASD. The 230 charts with high evidence of ASD were selected to undergo the rigorous review process where we identified sleep problems as described below and in Figure 1.

**Figure 1.**
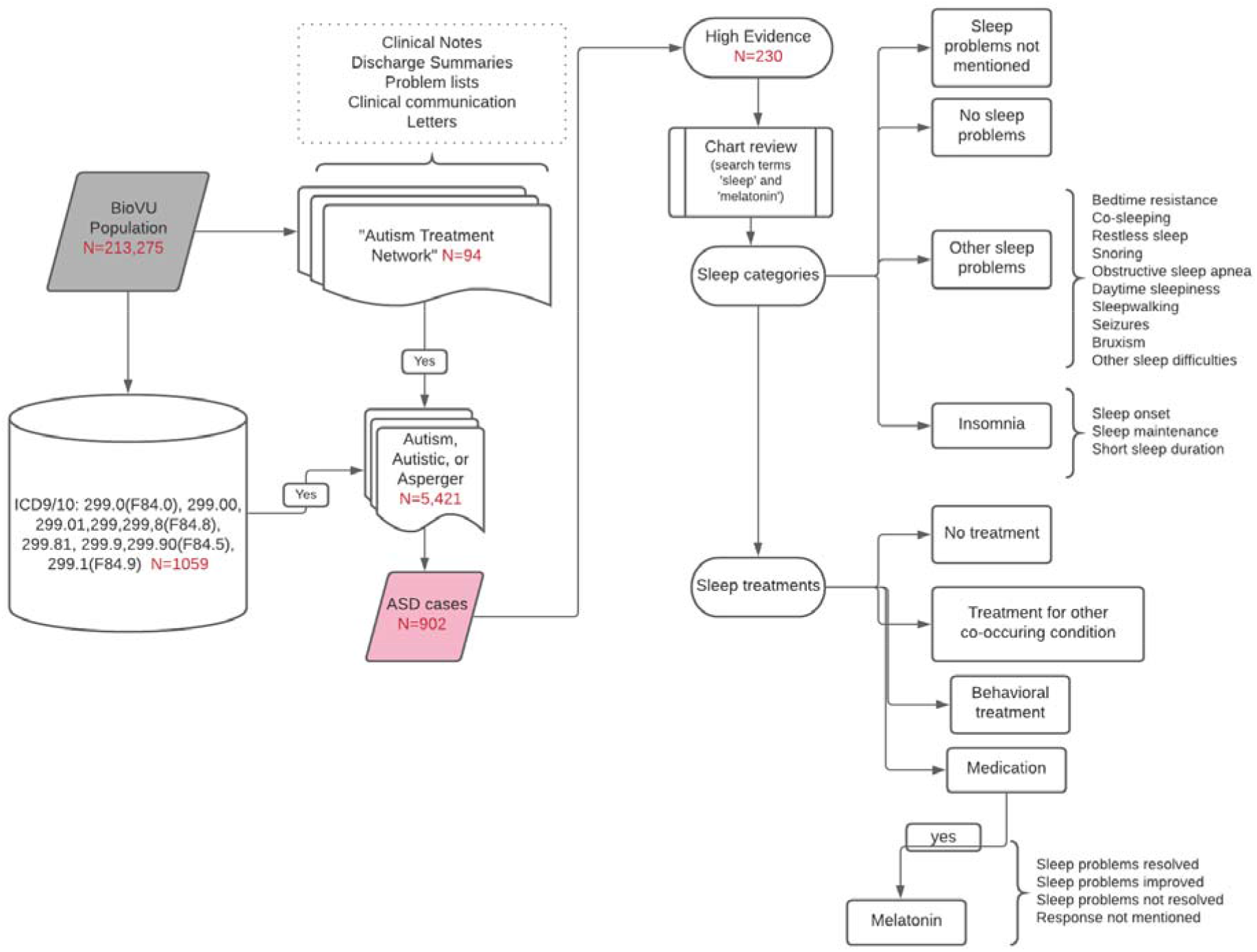
Identification of individuals with ASD and their sleep problems and treatments

### Identification of sleep problems in individuals with autism

#### Pilot Study

Initially, three reviewers (EVS, MN, BAM) each extensively reviewed 10 charts separately and then met to decide on the best strategy to be used to adequately capture the types of sleep problems reported in the EHR. This process allowed us to establish search terms to be used as well as categories for the types of sleep and specific sleep problems to be documented. A REDCap data form was created that was then used to track the required information of each chart. REDCap forms are a HIPPA-compliant database that could be access by different individuals at once. To further confirm that we were being inclusive of all sleep problems, each reviewer examined an additional 15 charts with our defined process and again met to analyze them. In this way, we were able to carefully review and then test this process using a total of 25 charts in this pilot study.

#### Chart Review

Our chart review proceeded with two of the authors (EVS, MN) reviewing the charts separately and then comparing results to ensure agreement on the types of sleep problems and treatments present in each chart. The two reviewers examined between 20-50 charts at one time. Each reviewer documented reference quotes from the chart to support their choices. Once the two reviewers met if there was disagreement then they would first try to rectify the disagreement by referencing the quotes as supporting evidence. If the disagreement remained, then those charts were sent to a third reviewer. Our third reviewer was a sleep medicine physician with autism experience (BAM) who reviewed the responses and adjudicated the designation. With this method we reviewed 230 charts including those completed during the pilot study.

#### Defining Sleep Categories

Charts were reviewed using the search terms “sleep” and “melatonin”. Sleep problems were categorized into: (a) sleep onset insomnia; (b) sleep maintenance insomnia; (c) bedtime resistance; (d) short sleep duration; (e) co-sleeping; (f) restless sleep; (g) snoring; (h) obstructive sleep apnea; (i) daytime sleepiness; (j) sleepwalking; (k) seizures; (l) bruxism and (m) sleep difficulties without further detail. To fit into a category the chart reviewer used certain definitions. For example, to qualify as short sleep duration the patient needed to sleep for less than 6 hours per night (Itani et al., 2017). To qualify for sleep onset insomnia the patient had to take greater than 30 minutes to fall asleep (Smits et al., 2001). For daytime sleepiness, if the sleepiness was secondary to a medical condition or a medication side effect then it was not documented. For obstructive sleep apnea, it was documented if there was a clinical diagnosis of obstructive sleep apnea (OSA) or sleep disordered breathing (SDB) made by pediatric otolaryngology, if they underwent a tonsillectomy and adenoidectomy for the diagnosis of OSA or SDB, or if there was documentation of the patient undergoing a sleep study with results indicative of obstructive sleep apnea (i.e. apnea-hypopnea index of >1) (Roland et al., 2011). Co-sleeping was considered a sleep problem if it was co-sleeping outside of infancy. While co-sleeping can be a cultural tradition, several studies support the presence of co-sleeping to be related to sleep problems in both typically developing children and children with neurodevelopmental disabilities. Köse et al. (2017) found that co-sleeping with a parent increases the risk of sleep disorder in the child thirteen-fold. Another study of children with ASD found correlation between sleep problems and co-sleeping (Liu et al., 2006). It was documented if the patient had some type of insomnia (sleep onset, sleep maintenance or short sleep duration) and if the patient had other sleep problems, excluding insomnia. Patients were included if they had a sleep problem at any time within the entirety of their EHR. It was also documented if patients had no sleep problems or if there was no mention of sleep in the record.

#### Sleep Treatments

If the individual reported sleep problems, we would then seek to identify whether the patient received treatment and what type of treatment the individual received. To be more specific, sleep treatments were classified as (a) medication, (b) behavioral treatment, (c) none documented or (d) other medical/psychiatric conditions treated with expected effects on the patient’s sleep. This last category was created to acknowledge the presence of a co-occurring medical or psychiatric condition that was treated with medications that were either selected or expected to influence the patient’s sleep. Detailed notes were kept on the medications or behavioral treatments that were tried and the date and document name were recorded. It was documented whether the patient had received supplemental melatonin and the response they had to melatonin. The patient’s response was categorized into: (a) sleep problems resolved; (b) sleep problems improved but not completely resolved; (c) minimal improvement or no response; and (d) response not mentioned. Occasionally, the response to melatonin was documented as evolving over time. This evolution was noted and after all available clinical documents were reviewed a decision about the overall response to melatonin was made and placed into one of the four categories. If there was disagreement then adjudication as described above in the chart review process would occur.

#### ICD codes

We identified individuals from the 230 charts with high evidence of ASD who had at least one sleep-related ICD-9 and ICD-10 code, to examine whether the chart review method used (ICD codes vs. review of charts for key words) yielded different results.

## Results

### Description of High Evidence ASD Database

Of the 230 patients with high evidence of ASD, 178 (77.4%) were male and 52 (22.6%) were female, consistent with ASD being more common in males. The age range of the patients at the time of our chart review was 6-30 years of age with a mean age of 15.4 years (standard deviation 6.6 years). It was not possible to identify the age of the patient when sleep problems were first mentioned but most patient charts went back into childhood. Patient demographics are presented in Table 2. The majority of individuals had sleep problems (n= 197; 85.6%) with insomnia being the most commonly reported at 85.2% (n=168) of the patients with sleep problems. About a quarter (25.9%) of patients with ASD and any sleep problem were seen at least once in a sleep medicine clinic.

**Table 2:** Demographics of high evidence ASD database

| Demographics |  |  |
| --- | --- | --- |
| Age | 15.4 years (SD 6.6 yrs) |  |
| Sex |  | % |
|  | Male | 77.4 |
|  | Female | 22.6 |
| Race |  | % |
|  | White | 84.3 |
|  | American Indian or Alaska Native | 0.4 |
|  | Asian | 3 |
|  | Black or African American | 8.7 |
|  | Native Hawaiian or Other Pacific Islander | 0.4 |
|  | Other | 0.4 |
|  | Not available | 2.2 |
| Ethnicity |  | % |
|  | Hispanic or Latino | 7.4 |
|  | Not Hispanic or Latino | 90.9 |
|  | Not available | 1.3 |

### Insomnia Types

In the children and young adults that had insomnia, 72.6% had sleep onset insomnia, 81% had sleep maintenance insomnia, and 36.3% had short sleep duration. The three types of insomnia (sleep onset, sleep maintenance and short sleep duration) commonly overlapped. In 46 cases (27.4%) all three types of insomnia were found. In 50 cases (29.8%) sleep maintenance insomnia and sleep onset insomnia were co-existing. It was rarer for sleep onset insomnia and short sleep duration to co-exist with only two (1.2%) cases having those two types of insomnia. This was also true for sleep maintenance insomnia and short sleep duration. Six (or 3.6%) cases had only these two types of insomnia. When looking at each type of insomnia in isolation, 34 (20%) patients with insomnia had only sleep maintenance insomnia. In those individuals the most common coexisting sleep problem was snoring with 52.9% (n=18). In individuals with only sleep onset insomnia (n=24) the most common coexisting sleep problem was restless sleep in 45.8% (n=11). It was also more common for patients with sleep onset insomnia to have bedtime resistance than individuals with sleep maintenance insomnia (16.7% vs 5.9%). Lastly, just 7 patients had only short sleep duration.

Other sleep problems excluding insomnia were found in 29 patients (14.7% of patients with a sleep problem). For patients without insomnia, 13.8% had bedtime resistance, 7.9% had co-sleeping, 13.8% had restless sleep, 38% had snoring, 17.2% had OSA, 6.9% had daytime sleepiness, 0 patients had sleepwalking, 6.9% had seizures, 3.4% had bruxism and 34.5% had sleep difficulties not further specified.

For all patients with a sleep problem including ones with co-existing insomnia, 16.5% had bedtime resistance, 29.5% had co-sleeping, 33.5% had restless sleep, 43.5% had snoring, 20% had OSA, 26% had daytime sleepiness, 6.5% had parasomnias, 4% had seizures, 8% had bruxism and 5% had sleep difficulties not further specified. When comparing the existence of other sleep problems in patients with and without insomnia, a higher proportion of patients with insomnia had coexisting daytime sleepiness, restless sleep and co-sleeping. The highest prevalence of insomnia with co-sleeping was short sleep duration. The types of sleep problems with and without insomnia are compared in Figure 2.

**Figure 2.**
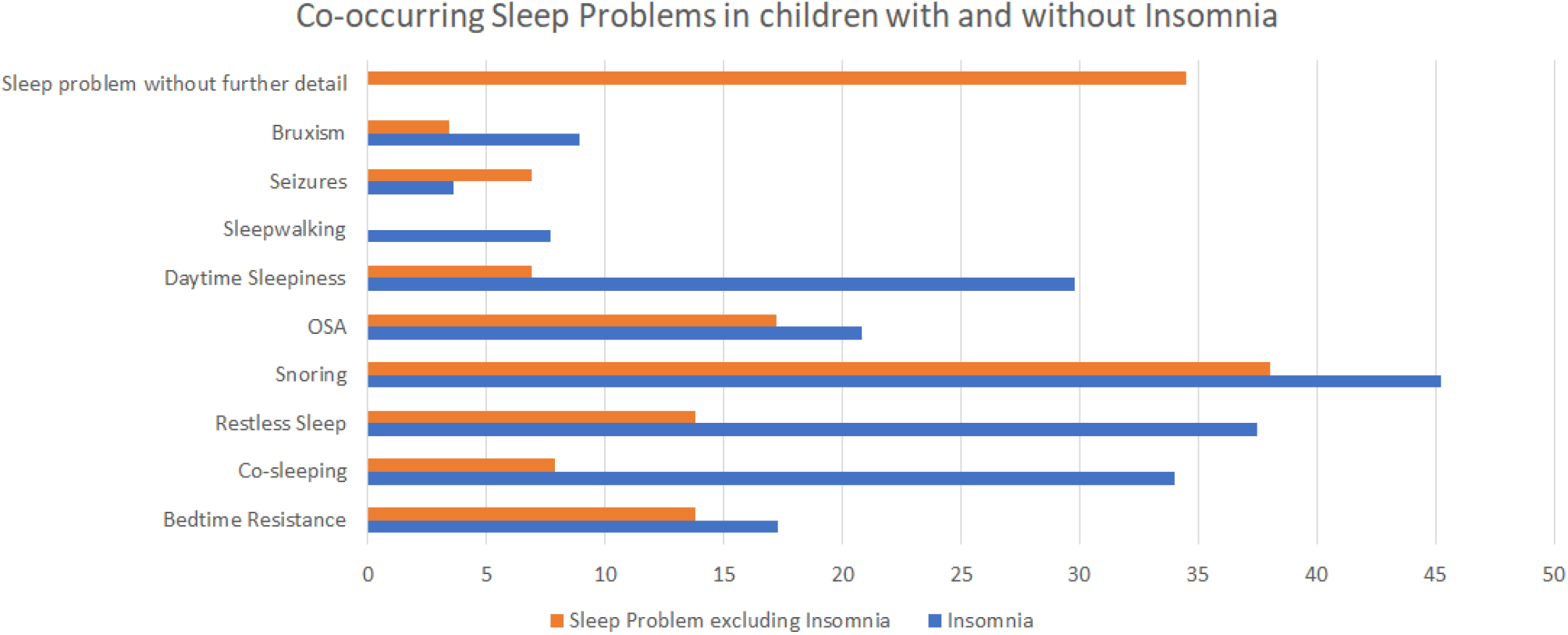
Co-occurring sleep problems in children with and without insomnia

Only 10.4% of patients were sleeping well and another 3.9% had no mention of sleep in their electronic health record.

In terms of treatments for disordered sleep, 138 patients (69%) received a medication for sleep. By far the most common medication for sleep was melatonin with 92% of the patients that received a medication getting melatonin. Behavioral treatments were documented as being used in 101 patients (50.5%). Eighty patients (40%) of patients with sleep problems received both a sleep medication and behavioral treatment. A treatment for a co-occurring medical condition that had the side effect of helping with sleep was documented in 45 patients (22.5%).

### Response to Supplemental Melatonin

Of the patient charts that had a documented response to melatonin, the largest percentage at 67% had some benefit to its initiation with either resolution of sleep problems (n=15, 18.1%) or sleep problems partially improved (n=41, 49.4%). Twenty-seven patients (32.5%) were documented to have no response to initiation of melatonin. Finally, 44 (35%) of the 127 patients with sleep problems and documented treatment with melatonin had no documentation in the chart on the response of melatonin. These patients commonly had melatonin on their medication list but no mention of taking the medication in their medical record.

### ICD codes versus chart review process

Table 3 shows the number of individuals identified through chart review versus the number of individuals identified by an ICD code in their medical record. Our results indicate that using the chart review process we were able to identify 50% more individuals with insomnia, and 68% more individuals with sleep problems other than insomnia, while the chart review process was also accurate in not finding a mention of sleep in the absence of an ICD code. There was one individual who had a sleep-related ICD code and where sleep was not mentioned specifically in this individual’s chart.

**Table 3.**
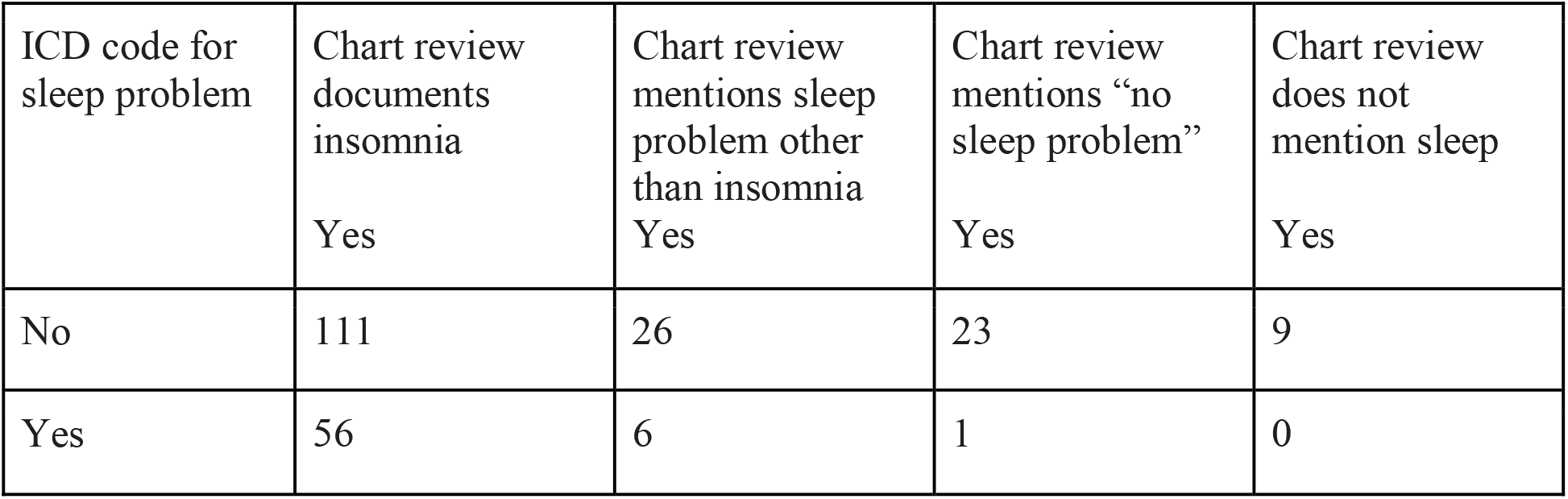
ICD code search for sleep problems compared to chart review

| ICD code for sleep problem | Chart review documents insomnia<br>Yes | Chart review mentions sleep problem other than insomnia<br>Yes | Chart review mentions "no sleep problem"<br>Yes | Chart review does not mention sleep<br>Yes |
| --- | --- | --- | --- | --- |
| No | 111 | 26 | 23 | 9 |
| Yes | 56 | 6 | 1 | 0 |

## Discussion

Our study provides a framework for characterizing sleep problems in ASD using de-identified patient records. We found similar rates of sleep problems in individuals with ASD demonstrated in prior studies. For example, Liu and colleagues found 86% of their cohort of 167 children had daily sleep problems. They reported higher rates of bedtime resistance than our study, but we found higher rates of insomnia (85% compared to 54%). These differences may be due to differences in the sleep categories and the ways sleep problems were defined in the respective studies.

A unique aspect of our study was the detail that was put into the chart review and categorization of sleep problems. Every chart was reviewed in its entirety by two separate reviewers for 13 unique sleep problems. Furthermore, sleep treatments, both medical and behavioral, were assessed with particular attention to melatonin and patients’ response to it. Charts commonly contained information about sleep problems and treatments over many years, thus allowing our review to assess whether a response to melatonin was short-lived or more permanent. To our knowledge this study is one of the first to utilize multiple facets of the EHR to characterize many different types of sleep problems.

We utilized a novel approach to the characterization of ASD within the EHR. Previous studies have utilized keyword searches as we utilized (Brooks et al., 2021). However, our algorithm further delineated the evidence of ASD in the chart as low, medium or high. These criteria allow for further certainty in a condition that can be challenging to diagnosis given its heterogenous presentation.

Our large study of 230 patients with high evidence of ASD were each evaluated with detailed reviews of sleep problems and treatment. In undertaking this review of medical records, we realized the importance of reviewing all notes and communications that were available in the EHR. As sleep problems can be brought up in many different settings and can change over time, this was necessary to get a complete understanding of the patient’s sleep issues. This method of thorough review was also helpful when trying to understand patients’ response to initiation of melatonin.

Overall, we found melatonin response to be challenging to discern in the EHR. A prior study looked at melatonin response in children with ASD and found 25% no longer had sleep concerns, 60% had improved sleep, 13% had no response and only 1% had an undetermined response (Andersen et al., 2008). In our review, for patients that had a documented response to melatonin, we found more patients that did not improve with melatonin-32.5% compared to only 13% in the Andersen study. This difference may be related to our higher percentage of patients with undocumented response to melatonin, which was 40% compared to only 1%. It is important to note that the Andersen study results were collected from one pediatrician’s clinical documentation and all patients were recommended behavioral treatments alongside melatonin administration. This may account for some of the differences in responsiveness of melatonin and for the lack of information about melatonin response. Finally, since melatonin is a dietary supplement, it was not always documented in the medication list of a patient but was mentioned in the chart that the patient was taking it.

During the review process, charts might report a patient was “sleeping well” but upon further review the patient would have melatonin on their medication list. To avoid contaminating the “sleeping well” control group, those charts would be marked as “other sleep problems” even though there was no explicit mention of a sleep problem.

Limitations of our design were the retrospective nature of the study and the challenges related to use of EHR data. Varied documentation by many different providers led to some difficulties acquiring consistent information. This led to missing data, for example, response to treatment, particularly behavioral treatments were often missing. While most charts had mention of sleep, there were a few charts that made no mention of sleep. It is possible those patients had sleep problems that were not asked about or documented.

Our study only looked at patients with high evidence of ASD. While this is an advantage in terms of confidence in their diagnosis, it may have caused ascertainment bias in terms of their sleep problems. All the patients with high evidence of ASD had documentation from physicians with expertise in ASD and may have been more likely to discuss sleep problems in that population. Finally, due to the lengthy review process, we limited our review to the high evidence for ASD charts.

## Conclusions

Our study can be used to guide future work using the EHR. We propose a method for differentiating ASD evidence and for evaluating challenging areas of study such as pediatric sleep problems. Much of what is in the literature is focused on sleep in children that were seen in a sleep clinic. Our study includes patients seen in many different clinical settings with about a quarter of them seen in a sleep clinic. Our approach to sleep problems in ASD improved the identification of patients with sleep problems that were identified in the EHR compared to only using ICD codes to identify patients with sleep problems.

In terms of feasibility, this study included a lengthy review process especially when compared to using ICD codes to identify patients. From this review process we were able to identify keywords that if used, could generate necessary information for determining sleep problems, sleep treatments and their responses. These keywords can be incorporated into future work with natural language processing to automate the process and allow for application on a larger scale.

## Data Availability

All data produced in the present work are contained in the manuscript

## List of Abbreviations

ASD: autism spectrum disorder
EHR: electronic health record
ICD: international classification of diseases
OSA: obstructive sleep apnea
PSG: polysomnography
SD: synthetic derivative
TRIAD: Treatment and Research Institute for Autism Spectrum Disorder

## Acknowledgements

*Synthetic Derivative and REDCap*

The project described was supported by the National Center for Research Resources, Grant UL1 RR024975-01, and is now at the National Center for Advancing Translational Sciences, Grant 2 UL1 TR000445-06. The content is solely the responsibility of the authors and does not necessarily represent the official views of the NIH.

*M. Niarchou*, The project was supported by Autism Speaks, Grant 11680

